# Reduced Plasma IL-40 and Total IgA Levels in Patients with Substance Use Disorders: Indicators of Impaired Humoral Immune Response

**DOI:** 10.1101/2025.04.10.25325594

**Authors:** Sheu Kadiri Rahamon, Abdulrahamon Adebayo Sikirullah, Sofiyat Opeyemi Bello, Oluremi Oladele, Victor Olufolahan Lasebikan

**Author notes:** **Corresponding Author:** Sheu Kadiri Rahamon, Department of Immunology, College of Medicine, University of Ibadan, Ibadan, Nigeria.

## Abstract

**Background:** Substance use disorders (SUDs) continue to be a public health challenge of significant importance. The immunomodulatory effects of substances of abuse have been extensively studied but there is a dearth of information on their effects on plasma interleukin-40 (IL-40) level, a biomarker of B cell activity, and its consequent effects on plasma total IgA level in patients with substance use disorders (SUD). Therefore, the plasma levels of IL-40 and total IgA in SUD patients were determined in this study.

**Methods:** Ninety adults comprising 50 SUD patients and 40 controls were enrolled into this case-control study. The SUD patients were stratified into groups based on the number of substances they abuse and plasma levels of IL-40 and IgA were determined using ELISA.

**Results:** Marijuana was the most abused substance (68.0%) and majority of the SUD patients (64.0%) were polydrug users. The median plasma IL-40 level was significantly lower in SUD patients compared with the controls. Similarly, the median plasma total IgA level was significantly lower in SUD patients compared with the controls. However, there were no significant differences in the plasma levels of IL-40 and IgA in SUD patients who abuse single substance, two substances, and three or more substances. The plasma IL-40 level had significant positive correlation with IgA in SUD patients.

**Conclusion:** Substance use disorder is associated with impaired humoral immune function, but the dysregulation appears not to be influenced by poly-drug use. Studies evaluating the mechanisms underlying humoral immune impairment in patients with substance use disorder and its potential clinical implications are suggested.

## Introduction

Substance use disorders (SUDs) are a global public health concern. Despite their associated morbidity and mortality, as well as existing drug laws, policies, and prevention strategies, the prevalence of drug and substance abuse continues to rise particularly among youths [1, 2]. In 2021, the estimated number of illegal drug users worldwide was around 296 million. Among these users, 39.5 million have a drug use disorder [3].

In Nigeria, drug and substance abuse is prevalent and remains a significant public health challenge. According to a 2018 report by the United Nations Office on Drugs and Crime (UNODC), the past-year prevalence of any drug use among individuals aged 15 to 64 years was 14.4%, representing approximately 14.3 million people. The report also indicated that among the six geopolitical zones, the South-West had the highest prevalence (22.4% or approximately 4.38 million users), followed by the South-South, South-East, North-East, North-West, and North-Central zones [4].

Drugs of abuse modulate the immune system primarily through receptor-mediated mechanisms; either by directly engaging receptors on immune cells or indirectly by interacting with analogous receptors in the nervous system [5]. They are associated with well-characterized changes in the levels of catecholamine such as dopamine, epinephrine, and norepinephrine that have both central and peripheral effects on neurotransmission and neuroendocrine signaling. Due to their activating effects on the sympathetic nervous system, abused stimulants may serve as potential mediators of the immune response. In particular, stimulant exposure is associated with disruption of the blood brain barrier and activation of the hypothalamic-pituitary-adrenal axis (HPA axis), which have consequences on immune function [6, 7].

Reports continue to show that patients with SUD have compromised immunity characterised by immunostimulation and immunosenescence [5, 8]. This immune dysfunction predisposes them to increased risk of infection requiring hospitalization or resulting in death [9]. Acute and chronic use of substances is associated with the dysregulation of the innate and adaptive immune response such as alterations in lymphocyte numbers, changes in cytokine expression and impairments in phagocytic functions [10-13]. In addition, studies have shown that immune factor signaling is associated with neural and behavioural aspects of addiction, such as drug seeking and resilience to relapse [14].

Inflammatory processes, especially the cell mediated immune response, play vital roles in the pathogenesis of neurological disorders associated with substance abuse [15]. However, dysfunction in humoral immune response in SUD patients continues to receive little attention. In 1976, Bogdal *et al*. [16] reported that there is an association between serum levels of immunoglobulins and alcohol use disorder (AUD). Similarly, elevated levels of immunoglobulin A (IgA) and IgE have been reported in SUD patients [15]. This alteration in immunoglobulin classes has also been reported in experimental studies [17]. Currently, there is lack of information on the dysregulation of cytokines involved in B cells homeostasis such as interleukin 40 (IL-40) in SUD patients.

IL-40, also known as chromosome 17 open reading frame 99 (*C17orf99*), is a B cell-associated cytokine implicated in humoral immune responses and B cell homeostasis [18-22]. An experimental study showed that it affects IgA production and, has direct influence on the composition of the intestinal microbiome in IL-40 knockout mice. In addition, the IL-40 knockout mice exhibited abnormalities in B cell populations, indicating the role of IL-40 in B cell development [18]. Although IL-40 may play a role in SUD-associated immune dysregulation, there is currently no information on its plasma level in SUD patients.

B cells count and subsets have been reported to be altered in patients with SUDs. Piepenbrink *et al*. [23] reported a significant, 2-fold increase in total B cells associated with increased activated B cell subsets in heroin injection drug users (IDU) compared with healthy controls. They also reported chronic B cell activation characterized by skewed plasma antibody profile with significant elevation in total IgM level and IgG3 and IgG4 subclasses but insignificantly different IgA level in heroin IDU. In addition, Wang *et al*. [24] reported that morphine significantly reduced IgA levels in animal models. Furthermore, Molina *et al*. [25] reported that chronic alcohol use resulted in decreased IgA secretion in the gut, contributing to systemic inflammation and liver damage. These reports clearly indicate that there is broad alteration in the steady-state humoral profile of patients with SUDs. Presently, information on the plasma IgA level in SUD patients is limited. Due to limited information on the plasma IL-40 and IgA levels in SUD patients, this study was conducted to determine the plasma levels of these biological markers in Nigerians with SUDs with a view to understanding how possible alteration in their plasma levels may impact the ability of SUD patients to generate optimal responses to infection.

## Materials and Methods

### Ethical Consideration

Ethical approval (UI/EC/24/0455) was obtained from the University of Ibadan/University College Hospital (UI/UCH) Joint Ethics Committee before the commencement of the study. Also, written informed consent was obtained from the study participants and where impossible, assent was obtained from the relatives or guardians.

### Study participants

A total of 90 participants consisting of 50 adults with substance use disorders (SUDs) and 40 apparently healthy adults who served as controls were enrolled into this case-control study using a convenient sampling method. SUD patients were enrolled from the Psychiatry Department, University College Hospital, Ibadan, Nigeria, and the New World Specialist Hospital, Ibadan. The controls were enrolled from the Ibadan metropolis and were certified free of any form of psychiatric disorder by a Consultant Psychiatrist. The study participants were recruited between 30th July, 2024 and 30th December, 2024.

### Sample size calculation

The sample size could not be calculated due to the lack of available reports on plasma levels of IL-40 in patients with SUDs, to the best of our knowledge, at the time this study was conducted. Therefore, a convenient sampling method was adopted and the study is thus, considered a preliminary study.

### Diagnosis of Substance Use Disorder

SUD was diagnosed using the Diagnostic and Statistical Manual of Mental Disorders, Fifth Edition (DSM-5) criteria [26].

### Exclusion criteria

Patients with history of schizophrenia, mood disorders and anxiety disorder were excluded from the study. Also, patients with history of autoimmune disorders and those on steroid therapy were excluded from the study.

### Data collection

Demographic data and clinical history of the study participants were obtained using a short-structured questionnaire.

### Blood sample collection

Venous blood samples (5 ml) was obtained from each study participant and dispensed into lithium heparinized sample bottles to obtain plasma which was stored at -20^0^C until analyzed.

### Laboratory Analysis

Plasma levels of IL-40 and IgA were determined using ELISA following the manufacturer’s instructions (Melsin Medical Co., China).

### Data Analysis

The Statistical Package for Social Sciences (SPSS), version 23.0 and GraphPad Prism, version 10.2.0 were used for data analysis. The data were assessed for Gaussian distribution using the Shapiro-Wilk test and Kolmogorov-Smirnov test. Thereafter, Mann Whitney *U* and Kruskal Wallis tests were used to determine differences in the median levels of IL-40 and IgA between two or more groups, respectively. Correlation between the variables was done using the Spearman rho correlation. *P-*values less than 0.05 (2-tailed) were considered as statistically significant. Results are presented as median (interquartile range).

## Results

The characteristics of the study participants are shown in the Table 1. Majority of the study participants were observed to be abusing marijuana (68.0%) and were polydrug abusers (64%).

**Table 1:** Characteristics of the study participants.

|  | SUD Patients (n = 50) | Controls (n = 40) |
| --- | --- | --- |
| Age | 28.86 ± 8.98 | 25.38 ± 6.01 |
| *Types of Substance abused* |  |  |
| <u>Marijuana</u> |  | - |
| Yes | 34 (68.0%) |  |
| No | 16 (32.0%) |  |
| <u>Cigarette</u> |  | - |
| Yes | 18 |  |
| No |  |  |
| <u>Alcohol</u> |  | - |
| Yes | 27 (54%) |  |
| No | 23 (46%) |  |
| <u>Tramadol</u> |  | - |
| Yes | 4 (8%) |  |
| No | 46 (92%) |  |
| <u>Heroin</u> |  | - |
| Yes | 2 (2%) |  |
| No | 48 (98%) |  |
| <u>Cocaine</u> |  | - |
| Yes | 3 (6%) |  |
| No | 47 (94%) |  |
| <u>Prescribed Drugs</u> |  | - |
| Yes | 1 (2%) |  |
| No | 49 (98%) |  |
| <u>Shisha</u> |  | - |
| Yes | 1 (2%) |  |
| No | 49 (98%) |  |
| <u>Codeine</u> |  | - |
| Yes | 5 (10%) |  |
| No | 45 (90%) |  |
| <u>Colorado</u> |  | - |
| Yes | 7 (14%) |  |
| No | 43 (86%)s |  |
| <u>Multiplicity of drug use</u> |  |  |
| Monodrug abusers | 18 (36%) | - |
| Polydrug abusers | 32 (64%) |  |

As shown in Figure 1, the median level of IL-40 was significantly lower in patients with SUD compared with the controls [146.02 pg/ml (100.51 – 237.78) vs 300.92 pg/ml (190.87 – 420.21), p-value = 0.000)]. Similarly, the median level of IgA was significantly lower in SUD patients compared with the controls [214.21 mg/ml (1370.27 – 3180.73) vs 5639.21 (3646.34 – 15577.83), P = 0.000)] (Figure 2).

**Figure 1.**
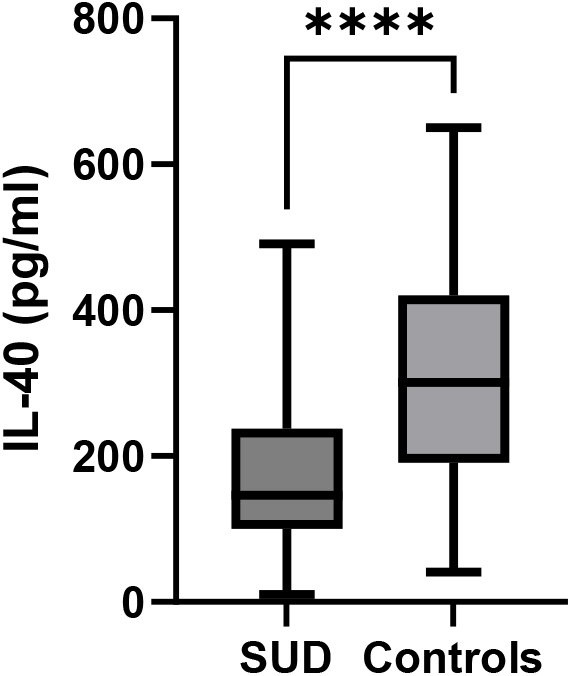
Plasma levels of interleukin-40 in SUD patients and controls

**Figure 2.**
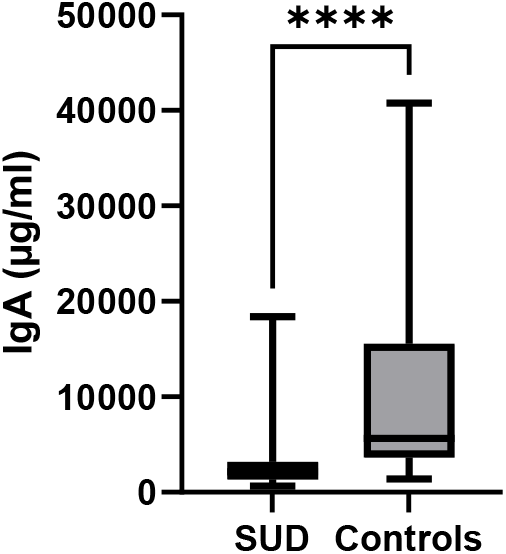
Plasma levels of immunoglobulin A in SUD patients and controls

Considering the number of substances abused, there were no significant differences in the median levels of IL-40 and IgA in patients monodrug abusers and polydrug abusers (Table 2).

**Table 2:** Plasma levels of IL-40 and IgA in monodrug and polydrug abusers.

| Parameters | Monodrug abusers<br>(n = 18) | Two substances<br>(n = 15) | Multiple substances<br>(n = 18) | P-value |
| --- | --- | --- | --- | --- |
| IL-40 (pg/ml) | 120.70<br>(77.53 - 166.70) | 181.30<br>(93.31 - 229.50) | 146.90<br>(104.20 - 324.10) | 0.262 |
| IgA (µg/ml) | 2526.49<br>(1491.59 - 4026.59) | 1825.48<br>(1352.28 - 2537.70) | 1996.50<br>(1185.81 - 3119.23) | 0.250 |

IL-40 = Interleukin-40, IgA = Immunoglobulin A

As shown in Table 3, plasma IL-40 level had significant positive correlation with IgA in SUD patients.

**Table 3:**
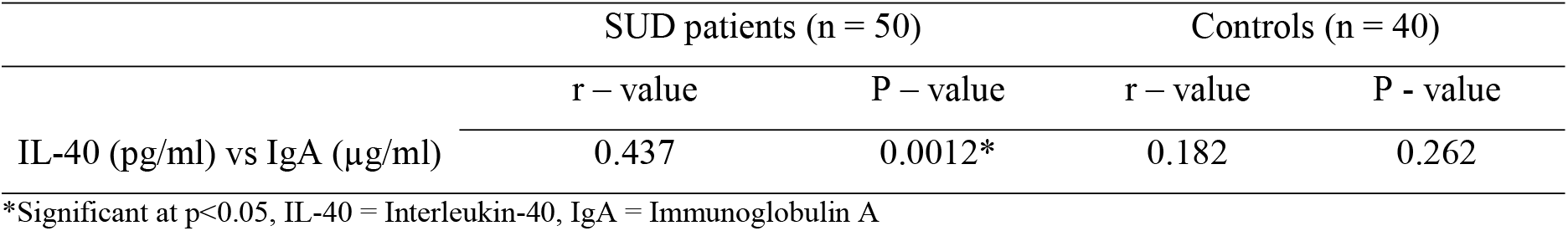
Correlation between the plasma levels of IL-40 and IgA in SUD patients and controls.

|  | SUD patients (n = 50) |  | Controls (n = 40) |  |
| --- | --- | --- | --- | --- |
|  | r – value | P – value | r – value | P - value |
| IL-40 (pg/ml) vs IgA (µg/ml) | 0.437 | 0.0012* | 0.182 | 0.262 |

\*Significant at p<0.05, IL-40 = Interleukin-40, IgA = Immunoglobulin A

## Discussion

Cannabis use disorder (CUD) is a global growing concern, with around 24% of individuals seeking treatment for SUD being diagnosed with the condition [27]. In 2010, it accounted for approximately 2 million disability-adjusted life years (DALYs) worldwide [28]. In this study, marijuana was observed to be the most abused substance in the SUD patients. This observation is in line with previous reports. In 2019, the World Drug Report showed that approximately 200 million people used cannabis, highlighting its widespread consumption [29]. Similarly, Morcuende *et al*. [30] reported that cannabis is the most commonly used illicit drug, worldwide. These reports, together with the observation from this study, showed that cannabis use is a serious public health problem and thus, addressing the rising use of cannabis and its associated health implications should remain a critical priority for public health interventions.

IL-40 is a cytokine involved in formation of B cells in the bone marrow and IgA production [31]. It is a proinflammatory cytokine whose expression is upregulated in conditions characterized by heightened immune activity. Reports have shown that it plays a crucial role in humoral immune responses and has been implicated in various autoimmune diseases, such as rheumatoid arthritis and Sjögren’s syndrome [21]. The observed significant reduction in median IL-40 level in SUD patients compared to controls suggests a potential link between serum IL-40 level and SUD pathophysiology. Our observation could not be compared with previous reports as currently, there is lack of information on IL-40 level in SUD patients. Immune system dysregulation, characterised by a state of peripheral inflammation and neuroinflammation or immunosuppression, is a common feature in SUD patients and plays crucial roles in the course of the disorder including drug dependence and relapse [30, 32, 33]. The observed significantly low IL-40 levels observed in SUD patients may suggest an impaired immune response, potentially contributing to the chronic inflammation associated with the disorder. Alternatively, the observed significant reduction might be a consequence of various therapies for SUD. Navrátilová *et al*. [21] reported a decrease in IL-40 level following Rituximab therapy; a B cell depleting therapy. Therefore, evaluation of the plasma IL-40 level as a potential biomarker for response to therapy in SUD patients is suggested.

IgA is the second most abundant immunoglobulin type found in the body. It is the principal antibody protecting the mucosal surfaces in the gastrointestinal, respiratory, and genitourinary tracts [34, 35]. In this study, plasma total IgA level was significantly lower in SUD patients compared to controls. This finding is consistent with previous studies demonstrating that opioids and alcohol impair IgA production [36, 37]. Our observation could be due to the observed reduction in IL-40 level which resulted in downregualtion of IgA level. This is further buttressed by the observed significant positive correlation between IL-40 and IgA in SUD patients. The report of Roy and Loh [36] showed that morphine inhibits the production of cytokines that promote IgA secretion. This observation further highlights the immunosuppressive effects of substance abuse and could be one of the mechanisms likely contributing to the increased susceptibility of SUD patients to infections and likely poor response to vaccines. The report of Wang *et al*. [9] showed that fully vaccinated SUD patients had a significantly higher risk of COVID-19 breakthrough infections compared to those without SUDs. In addition, those with breakthrough infections were more likely to experience severe health outcomes, including hospitalization and death. Although the observed low IgA level is hypothesized to be a consequence of low IL-40 level, it could also be a consequence of the effects of therapy as hypergammaglobulinemia, particularly of IgA and IgE, has been reported in SUD patients [15]. Therefore, there is the need for further studies to understand the homeostasis of IL-40 in SUD patients and the possible effects of therapy on it with a view to clearly delineating the observed reduction in total IgA level in SUD patients.

Reports on the effects of poly drug abuse on immune functions are conflicting. Pacifici and colleagues [38] reported that combining opioids with other substances led to greater immune suppression in heroin and morphine-treated mice. However, Friedman *et al*. [32] reported that the specific type of drug abused often has a greater impact on immune functions than the number of substances abused. The observed lack of significant differences in the plasma levels of IL-40 and IgA in monodrug and polydrug abusers may indicate that drug use elicits similar immunomodulatory effects irrespective of the number of substances that are abused. This observation is in contrast with some studies that reported additive effects of polydrug use on immune dysfunction. The findings of Rahamon *et al*. [13] revealed elevated phagocytic activity in polydrug abusers compared to monodrug abusers and suggested that polydrug use has additive or synergistic effects on immune dysfunction. While there are no available reports on IL-40 and IgA levels in monodrug and polydrug abusers, observation from this study could suggest that there may be a threshold of immune suppression beyond which additional substances do not further reduce IL-40 or IgA levels. Roy and Loh [36] reported that opioids significantly suppressed IgA production and modulated cytokine levels, overshadowing any additional impact of other substances used. Based on the previous reports and the findings from this study, it could be suggested that polydrug abuse exerts skewed immunomodulatory effects on the innate and adaptive immune response.

It could be concluded that patients with substance use disorder exhibit impaired humoral immune function, as evidenced by significant reduction in IL-40 and IgA levels. However, this dysregulation does not appear to be influenced by the multiplicity of substances abused. Further research is recommended to elucidate the mechanisms underlying humoral immune impairment in SUD patients and to explore its potential clinical implications. Small sample size and inability to enroll drug naïve SUD patients are some of the limitations in this study.

## Data Availability

All data generated during this study are included in this published article

## Declarations

## Availability of data and materials

All data generated during this study are included in this published article.

## Authors’ contributions

SKR conceived and designed the study; SKR, AAS, SOB, OO and VOL collected the samples; AAS, SOB and SKR did the laboratory analysis, SKR wrote the initial draft, SKR, AAS, SOB, OO and VOL reviewed the final draft, SKR supervised the entire research.

## Acknowledgements

The authors would like to appreciate the nurses and other clinical staff of the New World Specialist Hospital, Ibadan and University College Hospital, Ibadan for their support during the study.

